# Perceptions of Complementary, Alternative, and Integrative Medicine: Insights from a Large-Scale International Cross-Sectional Survey of Surgery Researchers and Clinicians

**DOI:** 10.1101/2024.10.13.24315409

**Authors:** Jeremy Y. Ng, Brenda X. Lin, Holger Cramer

## Abstract

**Background:** Given the potential benefits of complementary, alternative, and integrative medicine (CAIM) in perioperative care and the high prevalence of their use alongside conventional treatments, understanding perceptions of CAIM during surgery is important.

**Methods:** A survey was conducted among authors who published in surgery journals. 40,074 clinicians and researchers were invited to participate. The survey included questions on demographics and CAIM perceptions.

**Results:** The survey received 599 responses, with most identifying as both researchers and clinicians (n=331, 55.3%). Mind-body therapies (n=212, 47.1%) were considered the most promising CAIM areas for surgery. Most respondents believed CAIM therapies are safe (n=184, 46.2%) but were uncertain about their effectiveness (n=153, 38.6% disagreed and n=169, 42.7% were neutral). Many agreed on the value of CAIM research (n=310, 77.9%), increased funding (n=224, 55.9%), and clinician training through formal (n=215, 52.9%) and supplementary (n=246, 61.8%) education.

**Conclusions:** Surgery clinicians and researchers show strong interest in more CAIM education and research. This study’s findings can guide the development of resources and training programs to improve CAIM knowledge and perceptions.

## Background

Surgery is a branch of medicine that is focused on treating injuries, illnesses, and disorders using manual and instrumental methods [1]. There are many surgical specialities, which include but are not limited to general surgery, plastic surgery, orthopaedic surgery, and thoracic surgery [1]. A large research focus in the field of surgery surrounds care during the perioperative period, which is defined as the time period between the contemplation of surgery and recovery [2]. The perioperative period encompasses the preoperative phase, the intraoperative phase, and the postoperative phase [3]. Many patients who undergo surgery experience anxiety in the preoperative and postoperative periods [4], which is directly correlated to a number of factors, including administration of anaesthetic agents, anxiety surrounding the surgery itself, or perceived postoperative pain [5]. Moreover, postoperative pain, stress, and anxiety frequently remains severe despite the drug treatment [5]. While often effective, there are also limitations to pharmaceutical medications, such as concerns about addiction to pain medication [6], negative side effects, and incomplete efficacy [7]. Research has also found that patients are interested in taking a more active role in their postoperative recovery and believe that personal attitudes (e.g., maintaining a positive mindset) and self-directed healing (e.g., through mind-body therapies or natural product usage) has a positive impact on recovery [8]. As a result, some patients are turning to complementary, alternative, and integrative medicine (CAIM) therapies to integrate more self-management in the healing process and supplement their conventional postoperative pain and recovery treatments [8]. Among patients across a range of surgical disciplines, the use of CAIM is widespread with studies indicating a prevalence of 23.0% to 65.9% to supplement their conventional perioperative care [9]. “Complementary medicine” can be defined as treatments utilized alongside conventional medicine, while “alternative medicine” replaces conventional medical approaches [10]. Examples of therapies that are used alternatively include Ayurveda and naturopathy [11]. In recent times, the field of integrative medicine has gained popularity by combining both conventional and complementary methods to offer a more comprehensive approach to healthcare [10,12,13]. In the context of this research, these approaches will all be labelled as CAIM. Studies have found that the use of aromatherapy, massage, and acupuncture helps patients relieve preoperative anxiety [14]. Additionally, it was found that the incorporation of natural products (e.g., vitamins and herbs) and mind-body therapies (e.g., meditation) alongside their prescribed treatments were beneficial to patient perceptions of postoperative recovery [8]. Due to the potential benefits of CAIM use in perioperative care and the high prevalence of patients who already use them to supplement their conventional treatments, it is important to contextualize how CAIM use during the perioperative period of surgery is perceived.

Different CAIM therapies are used by patients undergoing various subtypes of surgery. For example, patients who undergo orthopaedic and trauma surgery frequently use exercise therapies, mindfulness, and traditional Chinese medicine for pain management [15]. Common CAIM treatments that are generally used among patients who undergo surgery include herbal products, such as ginger chewing, and techniques, such as acupressure and aromatherapy for postoperative treatment of nausea and vomiting [16]. The widespread use of CAIM treatments has an impact on the periods surrounding surgery [17]. These treatments can carry potential risks, especially concerning significant side effects and adverse drug interactions, such as reduced blood clotting, when herbal medications are combined with conventional therapies in surgical care [18]. Furthermore, patients often withhold information about their CAIM use from conventional healthcare providers due to past negative experiences or fear of their providers’ preconceptions regarding CAIM [19]. Despite the potential adverse side effects of some CAIM therapies, several randomized control trials for aromatherapy [20] and massage therapy [21] have reported positive outcomes of CAIM interventions in perioperative care. Though some healthcare professionals see CAIM as a beneficial complement to traditional healthcare, others harbour doubts regarding its efficacy and safety [22]. Given the potentially serious complications associated with combining CAIM treatments with conventional medications, it is important to improve our comprehension of how surgical researchers and clinicians perceive CAIM. The perception of CAIM among surgery researchers remains largely unexplored, and there is a paucity of scholarly literature addressing this topic outside of researchers who possess a particular focus on CAIM [23]. Various studies have been conducted on the knowledge and perception of CAIM among surgery researchers and surgical care providers in Sweden [24,25] and Hungary [19]. To the best of our knowledge, no international studies collected the perceptions of surgical researchers and clinicians on CAIM.

This study seeks to investigate the perspectives of both surgical researchers and clinicians regarding CAIM through an international, cross-sectional survey study design. The findings from this survey have the potential to provide insight into the challenges and opportunities associated with the use of CAIM in perioperative care. Ultimately, this research may help to better understand surgery researchers’ and clinicians’ perspectives on CAIM, which can assist in the development of educational resources for CAIM use in surgery research and practice.

## Methods

### Transparency Statement

Ethics approval was granted to conduct this study from the Research Ethics Board at the University Hospital Tübingen (REB Number: 389/2023BO2). The study protocol was registered and made accessible on the Open Science Framework (OSF). The study materials and raw data can also be found on OSF. This manuscript is reported in accordance with the STROBE (Strengthening the Reporting of Observational Studies in Epidemiology) [26] cross-sectional design reporting guideline and the CHERRIES checklist (Checklist for Reporting Results of Internet E-Surveys) [27].

### Study Design

We conducted an anonymous, online, cross-sectional survey of all selected authors who have published in surgery journals indexed in Ovid MEDLINE in the past three years.

### Sampling Framework

A sample of corresponding authors who have published articles in various surgery journals between November 11, 2020 and October 15, 2023 were selected from a sample of general surgery journals as found on the National Library of Medicine (NLM) broad subjects (https://journal-reports.nlm.nih.gov/broad-subjects/). The NLM identifications (IDs) of the chosen journals were extracted. Subsequently, a search strategy was developed using these NLM IDs and used on Ovid MEDLINE. The resulting list of PubMed IDs (PMIDs) from this search was exported as a .csv file. An R script, constructed using the easyPubMed package [28], was executed to retrieve details such as authors’ names, affiliated institutions, and email addresses. All authors who have published manuscripts of any type were incorporated into this study. No power analysis has been included as this study is based on a convenience sample and is primarily focused on descriptive work without inferential testing. The full search strategy can be accessed through the following link: https://osf.io/ta3pg.

### Participant Recruitment

The corresponding authors of articles published in the selected surgery journals were contacted to participate in our survey. Only surgery researchers and clinicians were eligible to complete the closed survey. SurveyMonkey was used to dispatch emails to the authors included in our sample. These potential participants received an email generated from an authorized recruitment script, which included a comprehensive explanation of the study’s objectives, along with the survey link. Upon clicking the link, invitees were directed to read an informed consent form. Survey participants were asked to provide informed consent by clicking “Yes” to a question about consent agreement. After consenting, participants were then be guided to the first page containing survey questions.

Duplicate email addresses within our list of authors, corresponding with those who have published multiple manuscripts in our sample, were deduplicated prior to sending our recruitment emails. We projected a response rate of 5% to 10% from our pool of prospective participants. Reminder emails were sent to individuals at intervals of one, two, and three weeks following the initial invitation email, and the survey was closed four weeks after the final reminder email was sent out. No financial compensation was be provided for participation in this study, and respondents could skip any questions that they did not wish to answer.

### Survey Design

A survey was constructed and shared through the Survey Monkey platform. Participants were presented with an initial screening question, followed by a series of multiple-choice questions surrounding their demographic details. The rest of the survey gathered information about each participants’ perceptions of CAIM and its perceived benefits and challenges through a series of multiple-choice, multi-select, and Likert scale questions. At the end of the survey, participants had the opportunity to answer an open-ended question about any additional comments they may have. It was anticipated that the survey will take approximately 15 minutes to complete. A copy of the full survey can be accessed through the following link: https://osf.io/svhzy

### Data Management and Analysis

This study does not have formal hypotheses. Fundamental descriptive statistics, such as counts and percentages, were generated by analyzing the quantitative data using GraphPad Prism 9. A thematic content analysis was also performed on the qualitative data. One members of the research team individually coded the received responses. The final codes were systematically organized and presented in distinct tables for reporting purposes. The Checklist for Reporting Results of Internet E-Surveys (CHERRIES) was also used to guide the reporting of this survey [27].

## Results

### Search Results

Of the 40074 emails that were sent, 15981 were unopened and 4102 bounced back. The bounced emails were excluded from response rate calculations. There were 599 total survey responses (corresponding to a 1.5% response rate for opened and unopened emails and 3.1% response rate for opened emails), which is lower than the anticipated 5-10%. On average, the survey took 8 minutes and 44 seconds to complete and had a 67% completion rate. The raw, deidentified survey data can be accessed through this link: https://osf.io/6bma5

### Demographics

Of the survey respondents, most identified themselves as both clinicians and researchers (n=331, 55.3%), with some as solely researchers (n=131, 21.9%) and some as solely clinicians (n=51, 8.5%). Regarding geographical location according to the World Health Organization World Regions, most respondents were from Europe (n=212, 41.9%) and the Americas (n=133, 26.3%). Approximately half of respondents identified themselves as clinicians (n=255, 50.3%), with identification as a faculty member (n=243, 47.9%) closely following. Most respondents described themselves as senior researchers or clinicians (n=279, 55.0%), with over 10 years of starting their careers post formal education. Lastly, most respondents classified their primary research area to be clinical research (n=353, 69.6%). The full table regarding demographical information can be found in **Table 1**.

**Table 1:** Demographics of Survey Respondents.

|  |  |
| --- | --- |
| **Sex (n=506)** |  |
| Female | 139 (27.5%) |
| Male | 361 (71.3%) |
| Intersex | 1 (0.2%) |
| Prefer not to say | 4 (0.8%) |
| Prefer to self-describe | 1 (0.2%) |
| **Age (n=507)** |  |
| Under 18 | 0 (0%) |
| 18-24 | 4 (0.8%) |
| 25-34 | 92 (18.2%) |
| 35-44 | 179 (35.3%) |
| 45-54 | 113 (22.3%) |
| 55-64 | 73 (14.4%) |
| 65 or older | 41 (8.1%) |
| Prefer not to say | 5 (1%) |
| **Visible minority (n=505)** |  |
| Yes | 94 (18.6%) |
| No | 388 (76.8%) |
| Prefer not to say | 23 (4.6%) |
| **World Health Organization World Region (n=506)** |  |
| Africa | 26 (5.1%) |
| Americas | 133 (26.3%) |
| Eastern Mediterranean | 21 (4.2%) |
| Europe | 212 (41.9%) |
| South-East Asia | 84 (16.6%) |
| Western Pacific | 18 (3.6%) |
| Prefer not to say | 12 (2.4%) |
| **Profession (n=507)** |  |
| Clinician Student (e.g., medical student, nursing student, etc.) | 9 (1.8%) |
| Clinician (e.g., physician, nurse, etc.) | 255 (50.3%) |
| Graduate student | 25 (4.9%) |
| Postdoctoral fellow | 36 (7.1%) |
| Faculty member/Principal Investigator | 243 (47.9%) |
| Research support staff (E.g., research manager, research associate, technician) | 21 (4.1%) |
| Scientist in academia | 94 (18.5%) |
| Scientist in industry | 3 (0.6%) |
| Scientist in third sector (E.g., NGO, non-profit) | 6 (1.2%) |
| Government scientist | 13 (2.6%) |
| Other (please specify) | 23 (4.5%) |
| **Career stage (n=506)** |  |
| Graduate or clinician student | 26 (5.1%) |
| Early career researcher or clinician (<5 years of formally starting your career post formal education) | 76 (15%) |
| Mid-career researcher or clinician (5-10 years of starting your career post formal education) | 125 (24.7%) |
| Senior researcher or clinician (>10 years of starting your career post formal education) | 279 (55.1%) |
| **Primary research area (n=410)** |  |
| Clinical research | 353 (86.1%) |
| Preclinical research – in vivo | 59 (14.4%) |
| Preclinical research – in vitro | 47 (11.5%) |
| Health systems research | 53 (12.9%) |
| Health services research | 70 (17.1%) |
| Methods research | 49 (12%) |
| Epidemiological research | 59 (14.4%) |
| Other (please specify) | 21 (5.1%) |
| **Area of CAIM research (n=407)** |  |
| Mind-body therapies (e.g., meditation, biofeedback, hypnosis, yoga, tai chi, imagery, creative outlets) | 42 (10.3%) |
| Biologically based practices (e.g., vitamins and dietary supplements, botanicals, special foods and diets) | 76 (18.7%) |
| Manipulative and body-based practices (massage, chiropractic therapy, reflexology) | 29 (7.1%) |
| Biofield therapies (e.g., reiki, therapeutic touch) | 5 (1.2%) |
| Whole medical systems (e.g., Ayurvedic medicine, traditional Chinese medicine, acupuncture, homeopathy, naturopathic medicine) | 29 (7.1%) |
| I have never conducted any CAIM research | 282 (69.3%) |
| Other, please specify (e.g., methodology, CAIM professions, etc.) | 9 (2.2%) |

### Complementary, Alternative, and Integrative Medicine

Most respondents had never conducted any CAIM research (n=282, 69.3%). Mind-body therapies (n=212, 47.1%) and biologically based practices (n=204, 45.3%) were seen as the most promising areas of CAIM in surgery (**Fig. 1**). Many clinicians declared that they have had patients who have sought counselling or disclosed using several different CAIMs, with the most common being biologically based practices (n=230, 67.5%), followed by similar rates for whole medical systems (n=155, 45.5%), mind-body therapies (n=153, 44.9%), and manipulative and body-based practices (n=148, 43.4%). Most clinicians responded that only 0-10% of patients (n=159, 46.8%) disclosed or sought counselling on CAIM use in the past year. Many clinicians also responded that they have never practiced nor recommended CAIM to their patients (n=112, 32.84%). Most clinicians responded that they have never received formal (n=244, 72.0%) or supplemental (n=199, 58.7%) training in any areas of CAIM. Of the CAIM areas where clinicians have received training, the most common area was biologically based practices (n=50, 14.8% for formal training and n=74, 21.8% for supplemental training). About half of clinicians responded that they have been asked about CAIM occasionally (n=235, 52.2%) outside of a clinical setting. The overwhelming majority of survey respondents declared that they would use academic literature to seek additional information on CAIM (n=372, 82.7%).

**Figure 1:**
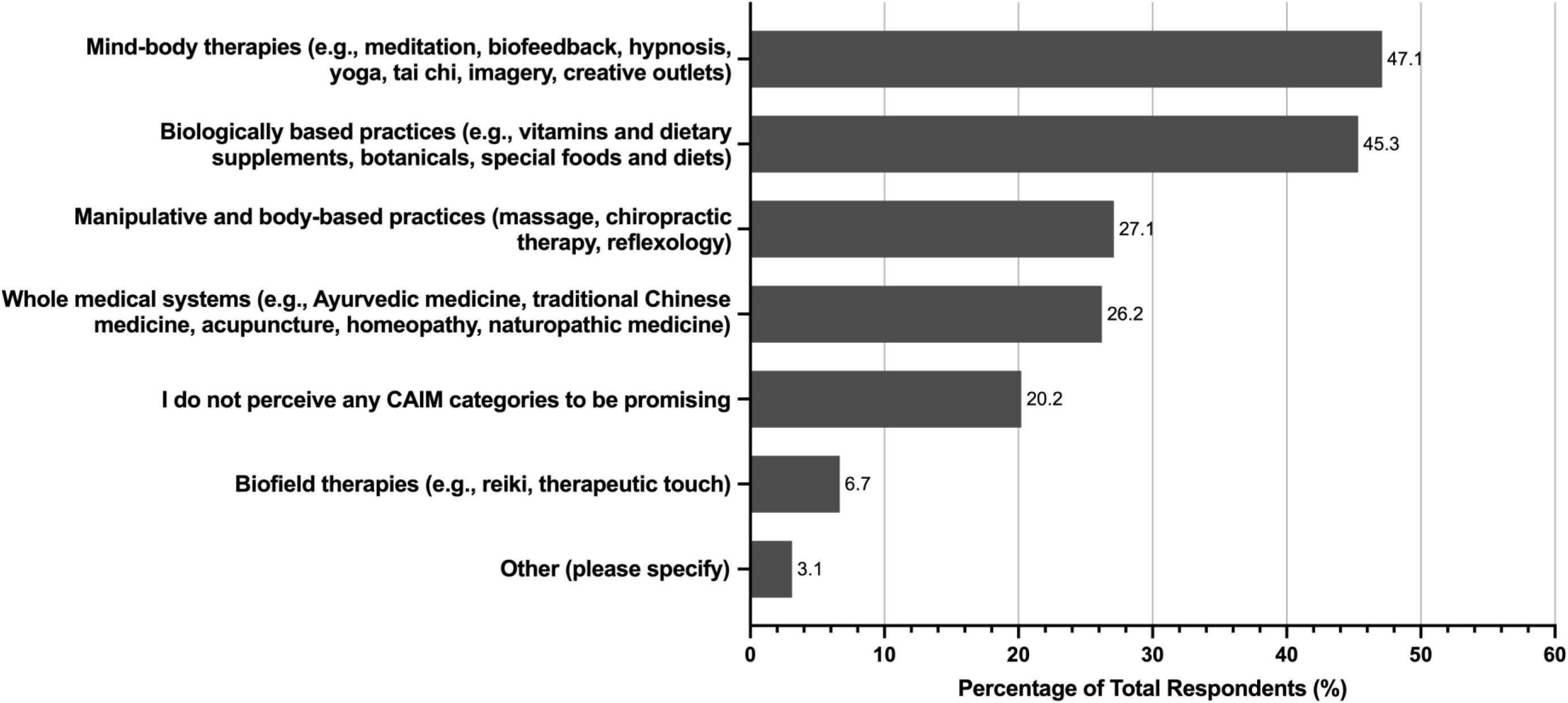
Areas of CAIM Seen by Respondents as the Most Promising for the Speciality of Surgery. CAIM: Complementary, Alternative, and Integrative Medicine.

The following responses about CAIM therapies were answered through a 5-point Likert scale, with the following options: “Strongly Disagree”, “Disagree”, “Neither Agree nor Disagree”, “Agree”, and “Strongly Agree”. When asked about the degree that respondents would agree with general CAIM statements, many agreed (n=156, 39.2%) or strongly agreed (n=28, 7.0%) that most CAIM therapies are safe, however, they were less confident that CAIM therapies are effective (n=153, 38.6% disagree and n=169, 42.7% neither agree nor disagree) (**Fig. 2**). Many respondents also either agreed or strongly agreed that there is value in conducting CAIM research (n=310, 77.9%), more funding should be allocated to CAIM research (n=224, 55.9%), and that clinicians should receive training on CAIM therapies through formal (n=215, 52.9%) and/or supplementary (n=246, 61.8%) education.

**Figure 2:**
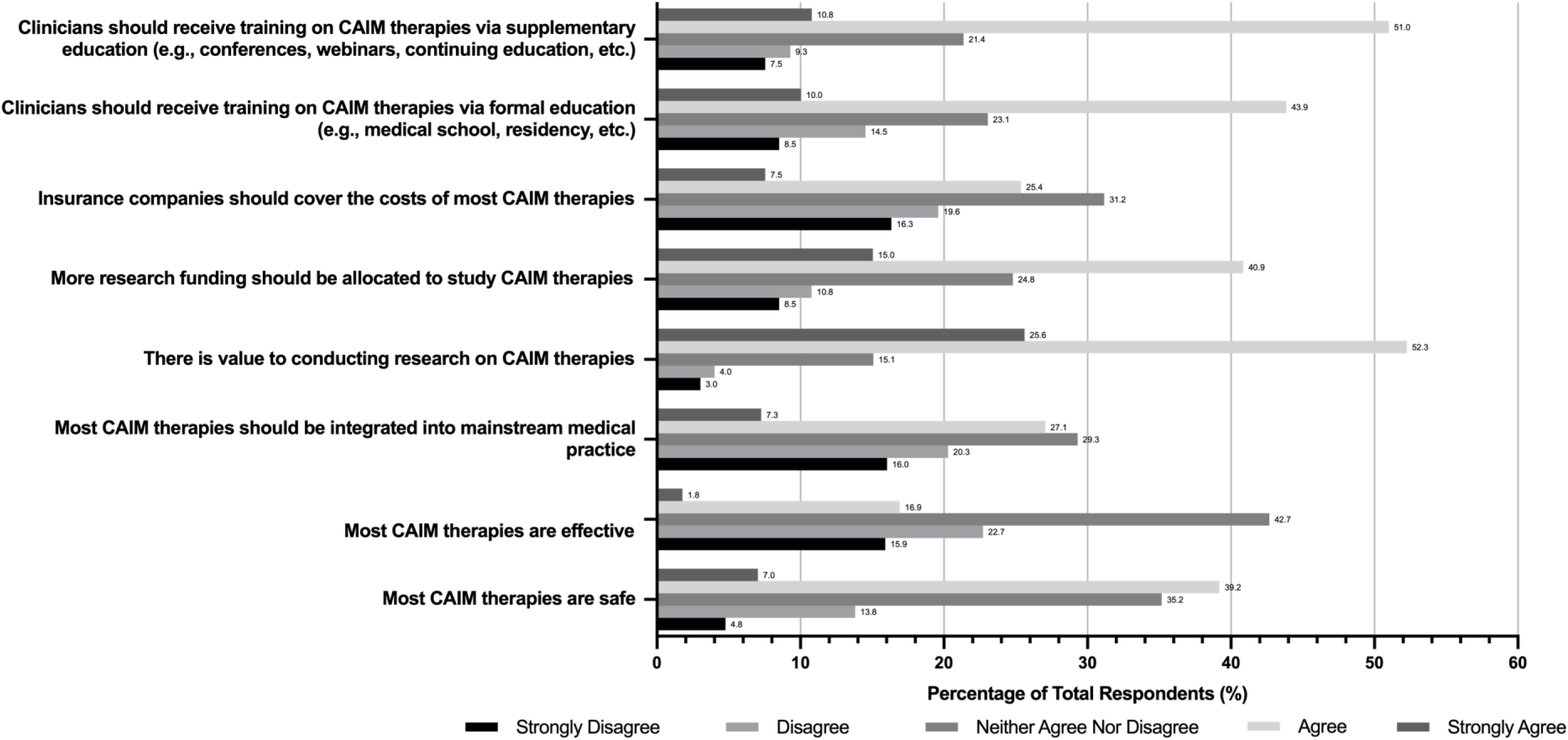
Degree of Agreement with general CAIM Statements. CAIM: Complementary, Alternative, and Integrative Medicine.

### CAIM Areas

Participants were then surveyed about the degree to which they agree with statements about mind-body therapies, biologically based therapies, manipulative and body therapies, biofield therapies, and whole medical systems (**Table 2**). Overall, the CAIM area that was perceived to be the most safe and effective was mind-body therapies, with 67% agreeing or strongly agreeing that they are safe and 35% agreeing that they are effective. In general, biofield therapies were perceived the least positively in response. Almost 30% of survey respondents disagreed or strongly disagreed that biofield therapies are effective. Similarly, while many respondents had diverse ranges of agreement to whether they believe different CAIM areas should be integrated into mainstream medicine, there was the least support for biofield therapies. Participants also saw the least value in conducting research, increasing research funding, and receiving both formal and supplementary education for biofield therapies compared to the other four CAIM areas.

**Table 2:**
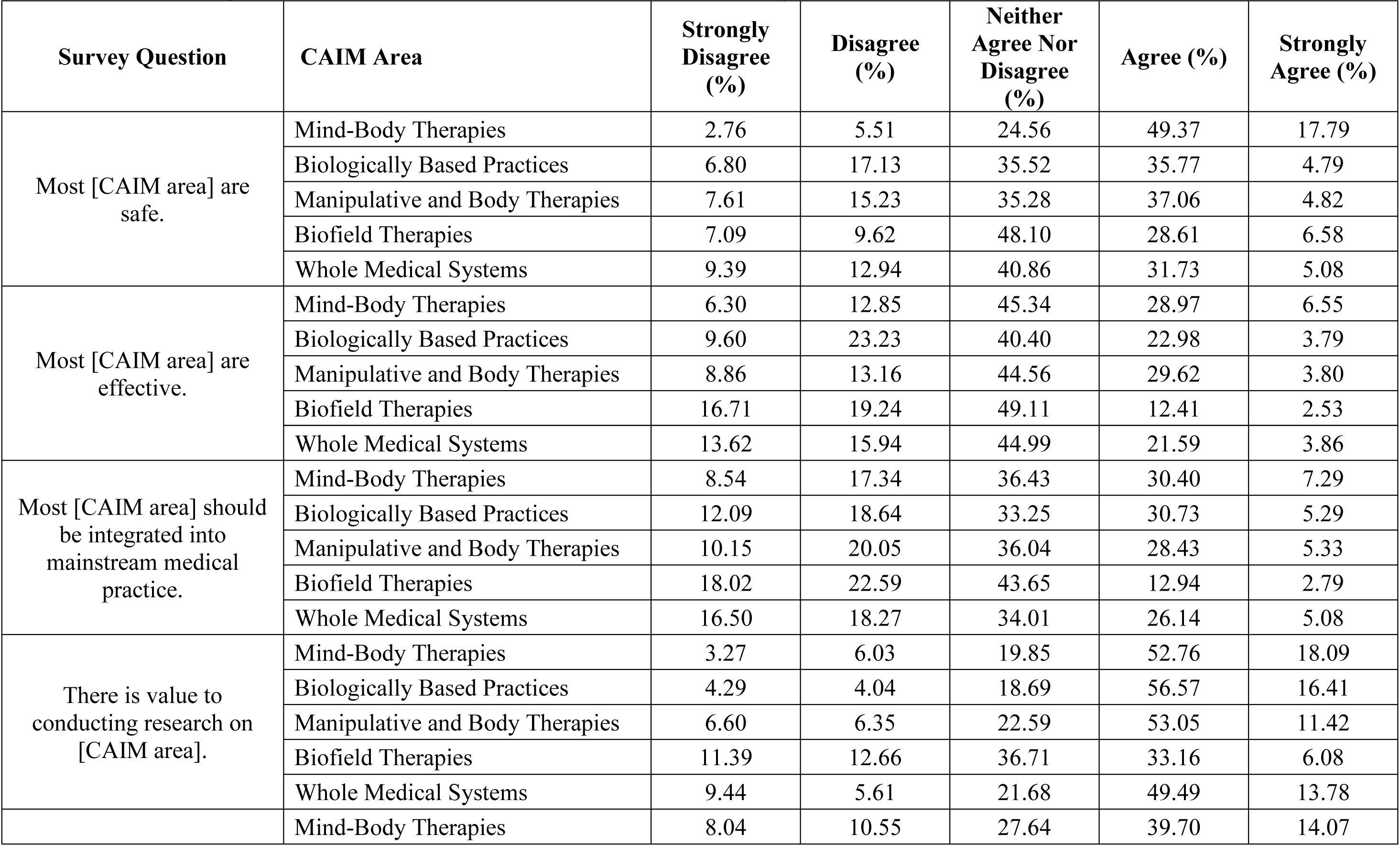
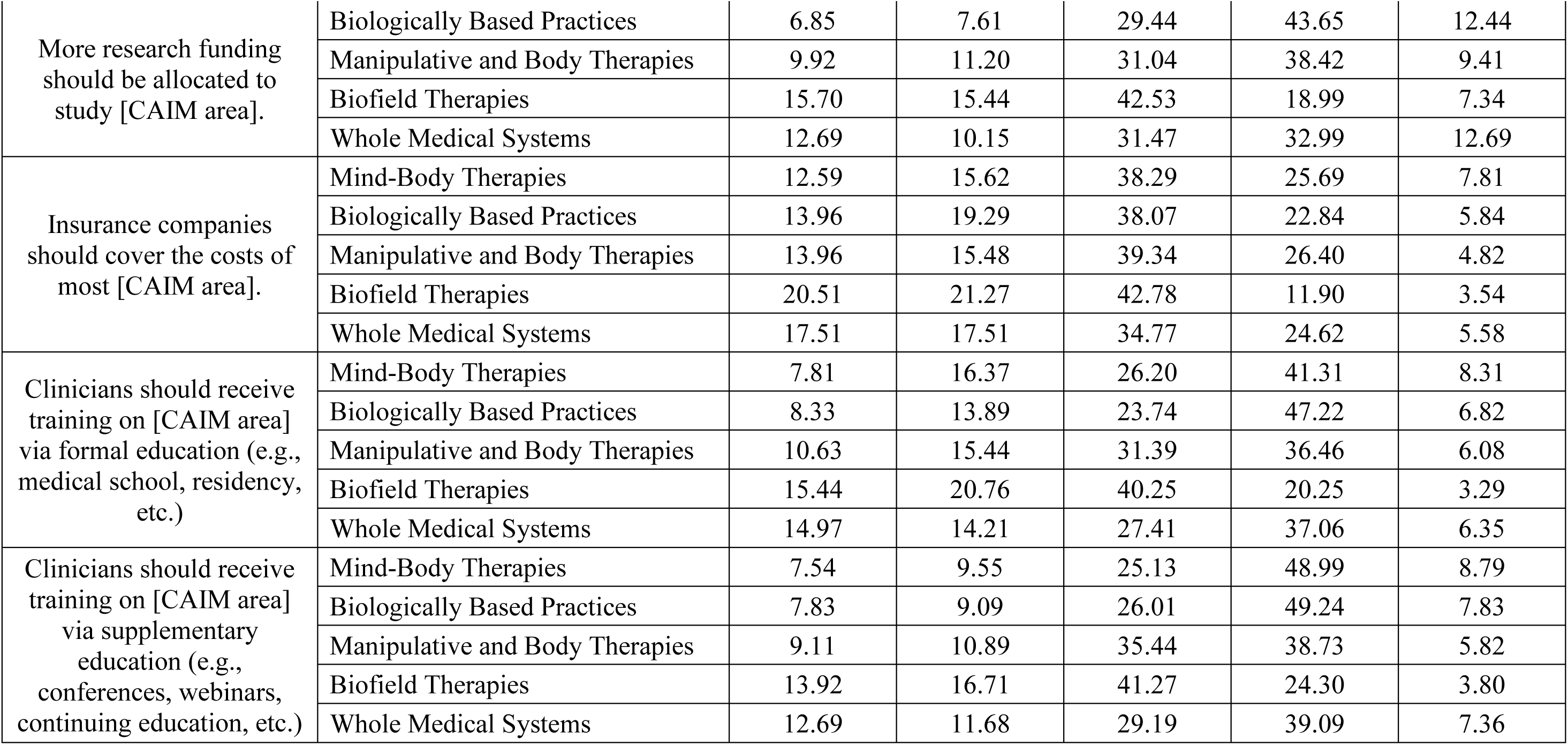
Degree to Which Respondents Agreed with Statements Regarding Mind-Body Therapies, Biologically Based Practices, Manipulative and Body Therapies, Biofield Therapies, and Whole Medical Systems.

### Comfort With Counselling About or Recommending CAIM

Overall, respondents were generally not comfortable with counselling about different CAIM therapies with their patients (**Fig. 3**). More clinicians were not comfortable with counselling (n=126, 41.2%) about CAIM therapies in general compared to clinicians who were (n=97, 31.7%). Survey respondents generally expressed discomfort with recommending CAIM therapies, with respondents being the most comfortable with recommending mind-body therapies (n=107, 37.5%) (**Fig. 4**). There was discomfort with recommending CAIM therapies in general (n=145, 47.4%) and clinicians were the least comfortable with recommending biofield therapies (n=163, 54.2%).

**Figure 3:**
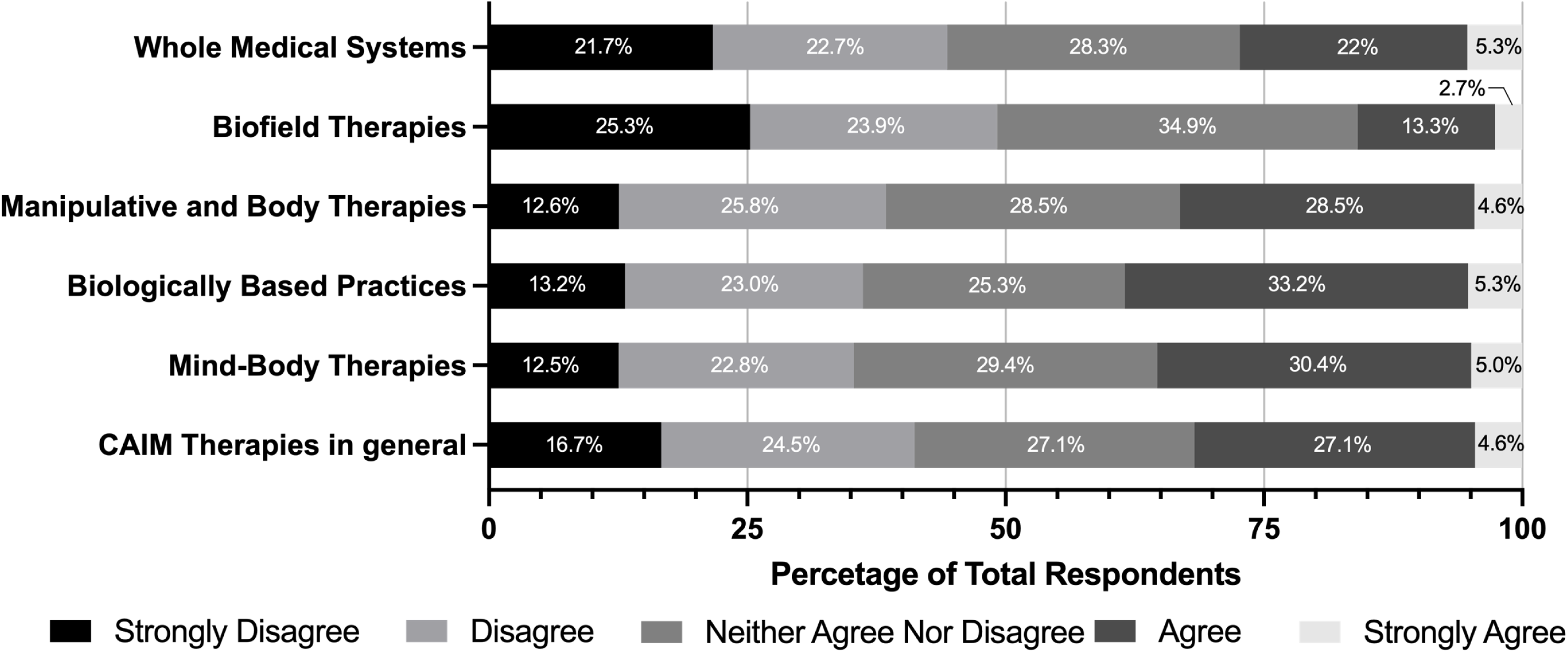
Degree of Comfort with Counselling Patients About CAIM Therapies. CAIM: Complementary, Alternative, and Integrative Medicine.

**Figure 4:**
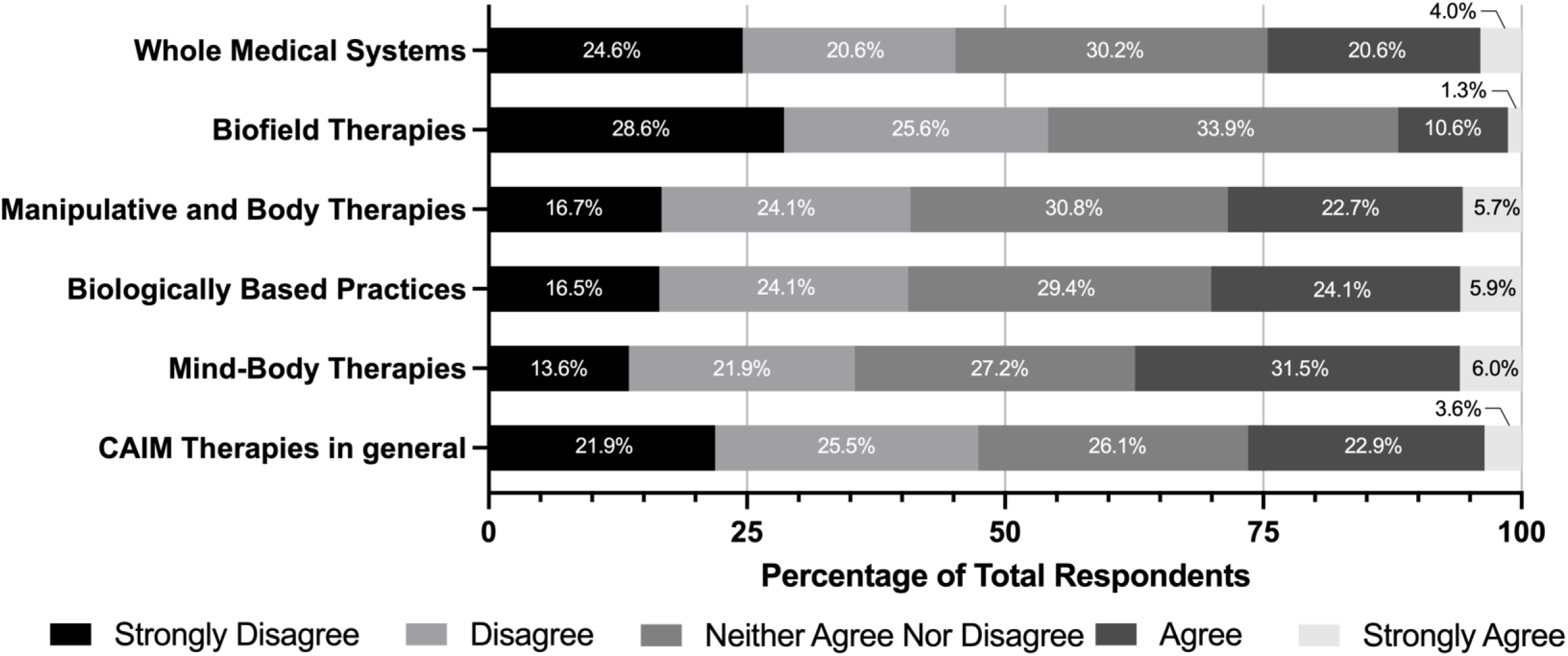
Degree of Comfort with Recommending CAIM Therapies to Patients. CAIM: Complementary, Alternative, and Integrative Medicine.

### Benefits and Challenges

The greatest benefits that respondents perceived to be associated with CAIM were expanded treatment options for patients (n=243, 61.8%), a focus on prevention and lifestyle changes (n=222, 56.5%), and a holistic approach to health and wellness (n=212, 52.9%) (**Fig. 5**). Conversely, the greatest challenges that respondents perceived to be associated with CAIM were a lack of scientific evidence for safety and efficacy (n=355, 88.8%), a lack of standardization in product quality and dosing (n=329, 82.3%), limited regulation and oversight (n=264, 66.0%), and difficulty in distinguishing legitimate practices from scams or fraudulent claims (n=255, 63.8%) (**Fig. 6**).

**Figure 5:**
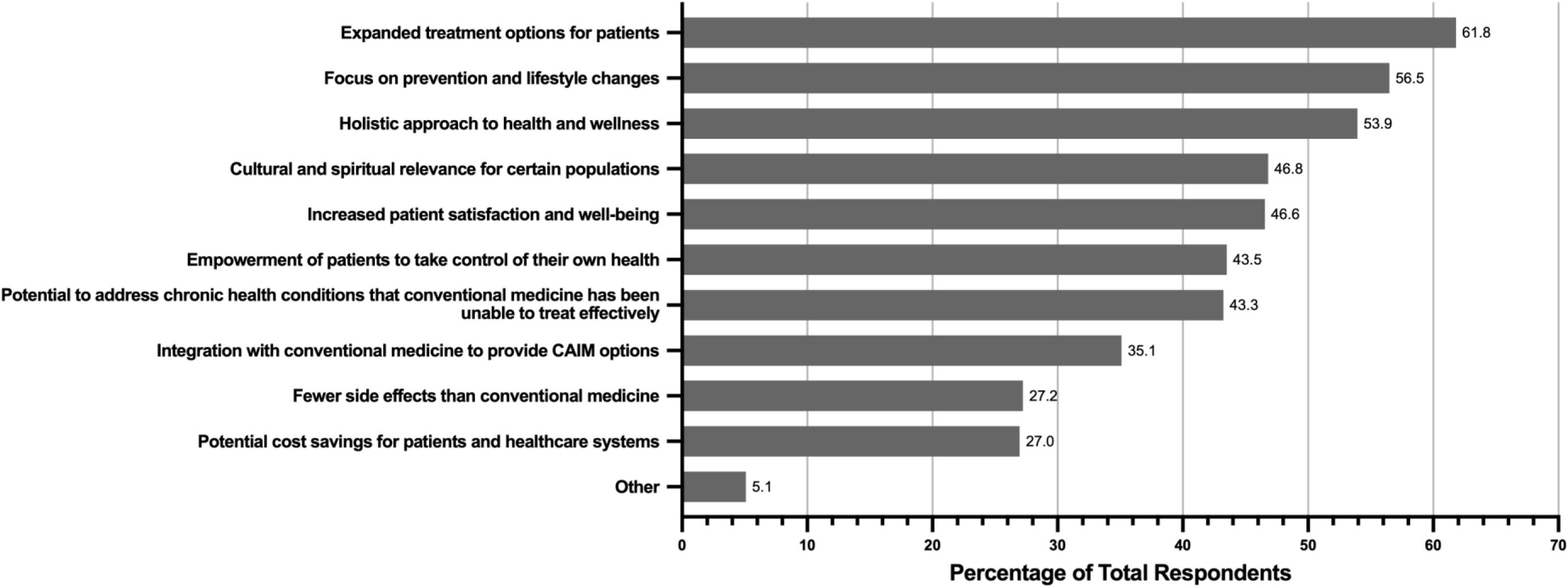
Perceived Benefits Associated with CAIM Within the Speciality of Surgery. CAIM: Complementary, Alternative, and Integrative Medicine.

**Figure 6:**
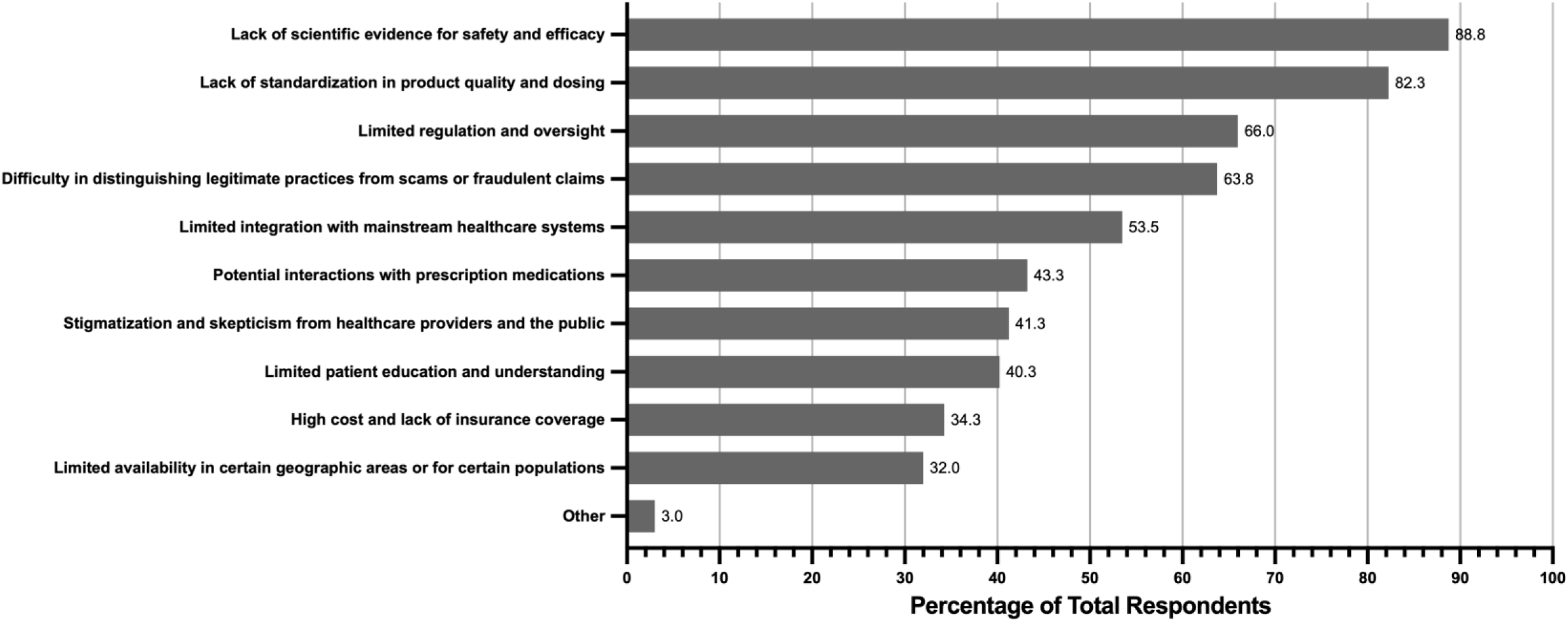
Perceived Challenges Associated with CAIM Within the Speciality of Surgery. CAIM: Complementary, Alternative, and Integrative Medicine.

### Thematic Analysis

In total, 22 codes were devised from the 63 open-ended responses. From these codes, 4 major themes were identified. The first was “benefits of CAIM”, which included four subthemes involving support of CAIM in general, support of specific CAIMs, and benefits for patients. The next theme was “integrative use of CAIM”, including two subthemes regarding how CAIM can be used alongside conventional treatments. Another theme was “concerns regarding the negative impacts of CAIM”, which included seven subthemes regarding the potentially predatory or fraudulent nature of some therapies, harm associated with insufficient training, a lack of standardization, and general opposition of its usage. The last theme was “insufficiency of CAIM research and education”, which included seven subthemes regarding a need for a greater evidence basis and patient or clinician education. The theme with the most coded responses was regarding the insufficiency of CAIM research and education. A copy of the full coding and thematic analysis data can be accessed through the following link: https://osf.io/pq3vt

## Discussion

The objective of this study was to investigate the existing perspectives of both surgical researchers and clinicians regarding CAIM. While various studies have been conducted on the knowledge and/or perception of CAIM among surgery researchers and surgical care providers in Sweden [24,25] and Hungary [19], or other scientific disciplines, to the best of our knowledge, there have been no international studies that have collected the perceptions of surgery clinicians and researchers on CAIM. Our findings are consistent with current studies within the literature. Studies have been conducted in the fields of psychiatry, oncology, and neurology that surveyed researchers and clinicians on their perceptions of CAIM. All three fields found very similar results regarding statements about different CAIM areas, with the most positive perceptions on CAIM research and education. Similarly to this study, the three fields also found mind-body therapies to be the most positively regarded [29–31]. However, while biologically based practices were found to be second most positively regarded for this study as well as the oncology and psychiatry studies, the neurology study had more negative perceptions of this CAIM area [29–31]. In a study performed on the perceptions of CAIM among Swedish surgery healthcare professionals, many interviewees mentioned concerns surrounding the evidence-basis of CAIM and the lack of knowledge surrounding the field [24]. Another study analysed the attitudes of CAIM among surgeons in Hungary, which yielded similar results regarding increased CAIM training through formal education [19]. As well, our results paralleled another study which found that surgery healthcare practitioners had more negative perceptions about biofield therapies and whole medical systems [25]. Compared to other types of CAIM treatments, the most research has been conducted on the attitudes of surgery clinicians on the use of acupuncture and acupressure in perioperative care. Similar to our study, these studies found that surgery clinicians had positive attitudes towards acupuncture and acupressure and supported its evidence-based usage, despite having little education on these therapies [32].

It was found that some areas of CAIM are more positively regarded than others, with the most positive perceptions of mind-body therapies and most negative perceptions of biofield therapies. Overall, there were greater positive perceptions of CAIM areas that included therapies that could be used complementary to conventional treatments (e.g., mind-body therapies) compared to those that are more frequently used alternatively to conventional treatments (e.g., whole medical systems). Mind-body therapies encompass treatments such as yoga, hypnosis, and meditation. This CAIM area may have more positive perceptions due a greater evidence basis and a lower risk of adverse interactions with conventional therapies. Several studies on the use of various mind-body therapies pre-operatively found that patients had lower levels of post-operative pain [33]. In contrast, biofield therapies may have more negative perceptions due to its comparatively lower evidence basis [34]. This lack of evidence-based research could also contribute to more the negative perceptions that surgery researchers and clinicians have on integrating CAIM therapies into mainstream medical practice. Additionally, though many survey respondents support CAIM research and increasing its funding, the open-ended responses also show that respondents believe CAIM research should only be funded or implemented if there is a realistic expectation for it to be beneficial. More specifically, studies on CAIM therapies should undergo the same methodological rigor as other areas of medicine to ensure their benefit for patients. Previous studies have shown that there is a high degree of heterogeneity in the quality of CAIM research, with some studies being more methodologically rigorous than others [23].

Though nearly half of all survey respondents agreed that most CAIM therapies are safe, less than a fifth agreed that they are effective. This may be due to a lack of evidence-based research confirming the benefits of CAIM. Some of the most common CAIM treatments used in surgery and perioperative care are massage therapy, herbal medicines, acupuncture, prayer, yoga, and relaxation therapy [35]. Within these treatments, there is substantially more evidence from methodologically sound studies for massage therapy, acupuncture, and yoga [21,36–38]. The thematic analysis also showed concerns surrounding the use of CAIM with conventional treatments. This presents a need for clinicians to be better educated on CAIM therapies as some therapies may have adverse effects with conventional medications that clinicians should be informed about prior to beginning surgery. Moreover, without CAIM training or education, some surgery clinicians may be dismissive of CAIM therapies despite there being evidence-based research supporting the safety and efficacy of some treatments. Half of clinicians reported that only 0 to 10% of patients disclosed the use of CAIM therapies within the past year. A previous study has shown that while the prevalence of CAIM use in cancer patients ranged from 11% to 95%, only 20% to 77% of these patients disclosed their use to their care providers [39]. There are diverse reasons for non-disclosure of CAIM therapies, which include lack of inquiry from medical providers, fear of provider disapproval, perception of disclosure as unimportant, and belief that providers lacked CAIM knowledge [40,41]. Though herbal medications remain a popular option among surgical patients [9], many studies have shown that non-disclosure of ingestible and natural CAIM therapies is more prevalent than non-disclosure of physical therapies [40,42]. This may be because the use of biologically based therapies (e.g., herbal supplements) is more discouraged as there is a greater risk for adverse interaction with conventional medical treatments [18].

While the majority of survey respondents have never had formal or supplemental training on CAIM, many agreed that clinicians should receive formal or supplemental training on most CAIM areas. This is consistent with researcher and physician attitudes from other studies on CAIM, where many clinicians or researchers believe that CAIM training and education is inadequate [43]. To better support patients’ interests in using CAIM therapies for reducing perioperative pain and anxiety, it is important for surgery clinicians to be educated on its usage and effects. One demographic that has been frequently targeted for CAIM education is medical students. Several studies have shown positive results when integrating CAIM into medical school curricula [44,45]. Moreover, strategies have also been developed to improve the integration of CAIM instruction into professional schools. For example, Mayo Clinic has created an integrative medicine education program for its medical students and physicians, using a combination of didactic and hands-on approaches [46]. CAIM education has also been added as a competency to the medical school curriculum in Switzerland [47]. Other strategies include greater top-down support for CAIM from institutional leaders, building relationships with reputable CAIM practitioners to develop instructional materials, as well as sustaining CAIM initiatives independent of financial support [48].

### Strengths and Limitations

Our study has several strengths. The cross-sectional survey design is time efficient, as the survey was administered on the SurveyMonkey platform and all data were collected at a single time point. Additionally, the international scope and selection of participants enhances the generalizability of our findings regarding surgery researcher perceptions of CAIM. Furthermore, participant contact information was obtained through NLM categorization, streamlining the process. As well, emails were only sent to researchers who have published within the last three years to minimize the chances of using invalid or inactive email addresses. Lastly, sending multiple reminders at one-week intervals with a one-month response window aims to improve participant response rates and potentially increase the sample size.

Regarding limitations, as the survey was administered in the English language, the result may not be representative of researchers and clinicians who are not English speakers. Moreover, because we used a sampling method that retrieves contact information from academic journals, we expected a higher response rate from researchers compared to clinicians. As well, due to our sampling method, our sample of clinicians is also not generalizable to clinicians who do not conduct research. We also anticipated a low survey participation rate due to various factors. These include the anonymity of participants to our research group, the unexpected nature of the survey invitations, and the likelihood that some participants may have changed their affiliations, lost access to their email, be on vacation, retired, or passed away. These factors could introduce nonresponse bias into our results. Moreover, our survey findings may be skewed toward individuals already familiar or have a positive attitude toward CAIM, as surgery researchers and clinicians with limited interest or negative perceptions of this area may be less inclined to participate. Lastly, participants could experience recall bias, where their responses may be affected by the differing accuracy and knowledge with which they remember their own experiences, given the survey’s reliance on self-reporting.

## Conclusions

This study investigated the knowledge and attitudes of surgery clinicians and researchers on CAIM. There were varying degrees of acceptance and attitudes across different CAIM areas, with mind-body therapies perceived to be the most promising and biofield therapies perceived to be the least. Our study provides valuable insight in understanding surgery clinician and researcher knowledge and perception of CAIM and establishes a need to improve CAIM training and education for these individuals. The information gathered from this study can be used as a foundation for creating resources and training programs that better inform researchers and clinicians about CAIM and address their knowledge gaps and barriers.

## Data Availability

All data and materials associated with this study have been posted on the Open Science Framework.

https://doi.org/10.17605/OSF.IO/8NUGQ

## List of Abbreviations

CAIM: complementary, alternative, and integrative medicine
CHERRIES: Checklist for Reporting Results of Internet E-Surveys
DOI: digital object identifier
MEDLINE: Medical Literature Analysis and Retrieval System Online
NLM: National Library of Medicine
OSF: Open Science Framework
PMID: PubMed identifier
REB: Research Ethics Board
STROBE: Strengthening the Reporting of Observational Studies in Epidemiology

## Declarations

### Ethics Approval and Consent to Participate

This study received approval from the University Tübingen Research Ethics board (REB Number: 389/2023BO2).

### Consent for Publication

All included study participants consented to participating in this study and to have their survey responses published in a peer reviewed journal.

### Availability of Data and Materials

All data and materials associated with this study have been posted on the Open Science Framework and can be found here: https://doi.org/10.17605/OSF.IO/8NUGQ

### Authors’ Contributions

JYN: designed and conceptualized the study, collected and analysed data, drafted the manuscript, and gave final approval of the version to be published.

BL: assisted with the collection and analysis of data, made critical revisions to the manuscript, and gave final approval of the version to be published.

HC: assisted with the analysis of data, made critical revisions to the manuscript, and gave final approval of the version to be published.

## References

[1] Definition of Surgery. AMA n.d. https://policysearch.ama-assn.org/policyfinder/detail/surgery?uri=%2FAMADoc%2FHOD.xml-0-4317.xml (accessed November 11, 2023).

[2] Davrieux CF, Pallermo M, Serra E, Houghton EJ, Acquafresca PA, Finger C, et al. Stages And Factors of The “Perioperative Process”: Points in Common With the Aeronautical Industry. Arquivos Brasileiros de Cirurgia Digestiva : ABCD 2019;32:e1423. 10.1590/0102-672020180001e1423.

[3] AORN. Resource Document for Non-Perioperative Staff Members Teaching Nursing Students about the Perioperative Setting. AORN; n.d.

[4] Attias S, Keinan Boker L, Arnon Z, Ben-Arye E, Bar’am A, Sroka G, et al. Effectiveness of Integrating Individualized and Generic Complementary Medicine Treatments with Standard Care Versus Standard Care Alone for Reducing Preoperative Anxiety. Journal of Clinical Anesthesia 2016;29:54–64. 10.1016/j.jclinane.2015.10.017.

[5] O’regan P, Wills T. The Growth of Complementary Therapies: And their Benefits in the Perioperative Setting. Journal of Perioperative Practice 2009;19:382–6. 10.1177/175045890901901102.

[6] Kallush A, Riley CA, Kacker A. Role of Complementary and Alternative Medicine in Otolaryngologic Perioperative Care. Ochsner Journal 2018;18:253–9. 10.31486/toj.18.0014.

[7] Reddy KK, Grossman L, Rogers GS. Common Complementary and Alternative Therapies with Potential Use in Dermatologic Surgery: Risks and Benefits. Journal of the American Academy of Dermatology 2013;68:e127–35. 10.1016/j.jaad.2011.06.030.

[8] Patel N, Pierson J, Lee T, Mast B, Lee BT, Estores I, et al. Utilization and Perception of Integrative Medicine Among Plastic Surgery Patients. Annals of Plastic Surgery 2017;78:557. 10.1097/SAP.0000000000000916.

[9] Yazici G, Erdogan Z, Bulut H, Ay A, Kalkan N, Atasayar S, et al. The Use of Complementary and Alternative Medicines Among Surgical Patients: A Survey Study. Journal of PeriAnesthesia Nursing 2019;34:322–9. 10.1016/j.jopan.2018.04.007.

[10] Complementary, Alternative, or Integrative Health: What’s In a Name? NCCIH n.d. https://www.nccih.nih.gov/health/complementary-alternative-or-integrative-health-whats-in-a-name (accessed July 5, 2024).

[11] Baars EW, Hamre HJ. Whole Medical Systems versus the System of Conventional Biomedicine: A Critical, Narrative Review of Similarities, Differences, and Factors That Promote the Integration Process. Evidence-Based Complementary and Alternative Medicine : eCAM 2017;2017. 10.1155/2017/4904930.

[12] Bell IR, Caspi O, Schwartz GER, Grant KL, Gaudet TW, Rychener D, et al. Integrative Medicine and Systemic Outcomes Research: Fpages in the Emergence of a New Model for Primary Health Care. Archives of Internal Medicine 2002;162:133–40. 10.1001/archinte.162.2.133.

[13] Gannotta R, Malik S, Chan AY, Urgun K, Hsu F, Vadera S. Integrative Medicine as a Vital Component of Patient Care. Cureus 2018;10. 10.7759/cureus.3098.

[14] Ebrahimi A, Eslami J, Darvishi I, Momeni K, Akbarzadeh M. Investigation of the Role of Complementary Medicine on Anxiety of Patients Before and After Surgery: A Review Study. Holistic Nursing Practice 2020;34:365–79. 10.1097/HNP.0000000000000414.

[15] Chou R, Deyo R, Friedly J, Skelly A, Hashimoto R, Weimer M, et al. Nonpharmacologic Therapies for Low Back Pain: A Systematic Review for an American College of Physicians Clinical Practice Guideline. Annals of Internal Medicine 2017;166:493–505. 10.7326/M16-2459.

[16] Department of Surgical Diseases Nursing, Istanbul University-Cerrahpasa, Graduate Education Institute, Istanbul, Turkey, Durust Sakalli G, Kara O, Department of Surgical Diseases Nursing, Istanbul University, Florence Nightingale Faculty of Nursing, Istanbul, Turkey. Use of Complementary and Integrative Methods in the Management of Postoperative Pain: A Narrative Literature Review. Mediterranean Nursing and Midwifery 2022;2:84–93. 10.5152/MNM.2022.222346.

[17] Waterbrook AL, Southall JC, Strout TD, Baumann MR. The Knowledge and Usage of Complementary and Alternative Medicine by Emergency Department Patients and Physicians. The Journal of Emergency Medicine 2010;39:569–75. 10.1016/j.jemermed.2008.01.007.

[18] Levy I, Attias S, Ben-Arye E, Goldstein L, Matter I, Somri M, et al. Perioperative Risks of Dietary and Herbal Supplements. World Journal of Surgery 2017;41:927–34. 10.1007/s00268-016-3825-2.

[19] Soós SÁ, Jeszenői N, Darvas K, Harsányi L. Complementary and alternative medicine: attitudes, knowledge and use among surgeons and anaesthesiologists in Hungary. BMC Complementary and Alternative Medicine 2016;16:443. 10.1186/s12906-016-1426-0.

[20] Huang H, Wang Q, Guan X, Zhang X, Kang J, Zhang Y, et al. Effect of Aromatherapy on Preoperative Anxiety in Adult Patients: A Meta-Analysis of Randomized Controlled Trials. Complementary Therapies in Clinical Practice 2021;42:101302. 10.1016/j.ctcp.2021.101302.

[21] Guo P-P, Fan S-L, Li P, Zhang X-H, Liu N, Wang J, et al. The Effectiveness of Massage on Peri-Operative Anxiety in Adults: A Meta-Analysis of Randomized Controlled Trials and Controlled Clinical Trials. Complementary Therapies in Clinical Practice 2020;41:101240. 10.1016/j.ctcp.2020.101240.

[22] Ng JY, Boon HS, Thompson AK, Whitehead CR. Making Sense of “Alternative”, “Complementary”, “Unconventional” and “Integrative” Medicine: Exploring the Terms and Meanings Through a Textual Analysis. BMC Complementary and Alternative Medicine 2016;16:134. 10.1186/s12906-016-1111-3.

[23] Polich G, Dole C, Kaptchuk TJ. The Need to Act a Little More ‘Scientific’: Biomedical Researchers Investigating Complementary and Alternative Medicine. Sociology of Health & Illness 2010;32:106–22. 10.1111/j.1467-9566.2009.01185.x.

[24] Bjerså K, Forsberg A, Fagevik Olsén M. Perceptions of Complementary Therapies Among Swedish Registered Professions in Surgical Care. Complementary Therapies in Clinical Practice 2011;17:44–9. 10.1016/j.ctcp.2010.05.004.

[25] Bjerså K, Stener Victorin E, Fagevik Olsén M. Knowledge About Complementary, Alternative and Integrative Medicine (CAM) Among Registered Health Care Providers in Swedish Surgical Care: A National Survey Among University Hospitals. BMC Complementary and Alternative Medicine 2012;12:42. 10.1186/1472-6882-12-42.

[26] Elm E von, Altman DG, Egger M, Pocock SJ, Gøtzsche PC, Vandenbroucke JP. The Strengthening the Reporting of Observational Studies in Epidemiology (STROBE) statement: guidelines for reporting observational studies. The Lancet 2007;370:1453–7. 10.1016/S0140-6736(07)61602-X.

[27] Eysenbach G. Improving the Quality of Web Surveys: The Checklist for Reporting Results of Internet E-Surveys (CHERRIES). Journal of Medical Internet Research 2004;6:e132. 10.2196/jmir.6.3.e34.

[28] Fantini D. easyPubMed: Search and Retrieve Scientific Publication Records from PubMed 2019.

[29] Ng JY, Kochhar J, Cramer H. Oncology researchers’ and clinicians’ perceptions of complementary, alternative, and integrative medicine: an international, cross-sectional survey. Supportive Care in Cancer 2024;32:615. 10.1007/s00520-024-08785-9.

[30] Ng JY, Kochhar J, Cramer H. An international, cross-sectional survey of psychiatry researchers and clinicians: perceptions of complementary, alternative, and integrative medicine. Frontiers in Psychiatry 2024;15. 10.3389/fpsyt.2024.1416803.

[31] Ng JY, Li SY, Cramer H. Perceptions and attitudes regarding complementary, alternative, and integrative medicine among published neurology authors: a large-scale, international cross-sectional survey. BMC Neurology 2024;24:215. 10.1186/s12883-024-03661-9.

[32] Zhang NM, Daly D, Terblanche M, Joshi S, Tacey M, Vesty G, et al. Doctors’ and Nurses’ Attitudes of Acupuncture and Acupressure use in Perioperative Care: An Australian National Survey. Pain Management Nursing 2022;23:800–10. 10.1016/j.pmn.2022.08.008.

[33] Hanley AW, Gililland J, Erickson J, Pelt C, Peters C, Rojas J, et al. Brief preoperative mind–body therapies for total joint arthroplasty patients: a randomized controlled trial. PAIN 2021;162:1749. 10.1097/j.pain.0000000000002195.

[34] Baldwin AL, Hammerschlag R. Biofield-based Therapies: A Systematic Review of Physiological Effects on Practitioners During Healing. EXPLORE 2014;10:150–61. 10.1016/j.explore.2014.02.003.

[35] Wu C, Weber W, Kozak L, Standish LJ, Ojemann JG, Ellenbogen RG, et al. A Survey of Complementary and Alternative Medicine (CAM) Awareness Among Neurosurgeons in Washington State. The Journal of Alternative and Complementary Medicine 2009;15:551–5. 10.1089/acm.2008.0427.

[36] Arruda APN, Zhang Y, Gomaa H, Bergamaschi C de C, Guimaraes CC, Righesso LAR, et al. Herbal medications for anxiety, depression, pain, nausea and vomiting related to preoperative surgical patients: a systematic review and meta-analysis of randomised controlled trials. BMJ Open 2019;9:e023729. 10.1136/bmjopen-2018-023729.

[37] Acar HV. Acupuncture and related techniques during perioperative period: A literature review. Complementary Therapies in Medicine 2016;29:48–55. 10.1016/j.ctim.2016.09.013.

[38] Azeez AM, Puri GD, Samra T, Singh M. Effect of Short-Term Yoga-Based-Breathing on Peri-Operative Anxiety in Patients Undergoing Cardiac Surgery. International Journal of Yoga 2021;14:163. 10.4103/ijoy.IJOY_120_20.

[39] Davis EL, Oh B, Butow PN, Mullan BA, Clarke S. Cancer Patient Disclosure and Patient-Doctor Communication of Complementary and Alternative Medicine Use: A Systematic Review. The Oncologist 2012;17:1475–81. 10.1634/theoncologist.2012-0223.

[40] Foley H, Steel A, Cramer H, Wardle J, Adams J. Disclosure of complementary medicine use to medical providers: a systematic review and meta-analysis. Scientific Reports 2019;9:1573. 10.1038/s41598-018-38279-8.

[41] Yukawa K, Ishikawa H, Yamazaki Y, Tsutani K, Kiuchi T. Patient health literacy and patient-physician communication regarding complementary and alternative medicine usage. European Journal of Integrative Medicine 2017;10:38–45. 10.1016/j.eujim.2017.02.003.

[42] Halpin SN, Potapragada NR, Bergquist SH, Jarrett T. Use and factors associated with non-disclosure of complementary and alternative medicine among older adults. Educational Gerontology 2020;46:18–25. 10.1080/03601277.2019.1698184.

[43] Patel SJ, Kemper KJ, Kitzmiller JP. Physician Perspectives on Education, Training, And Implementation of Complementary and Alternative Medicine. Advances in Medical Education and Practice 2017;8:499–503. 10.2147/AMEP.S138572.

[44] Ben-Arye E, Finkelstein A, Samuels N, Ben-Yehuda D, Schiff E, Reis S, et al. From skepticism to openness: a qualitative narrative analysis of medical students’ attitudes following an integrative medicine course. Supportive Care in Cancer 2022;30:4789–95. 10.1007/s00520-022-06888-9.

[45] Ahmed SM, Al-Mansour MA, Mohamed EY, Medani KA, Abdalla SM, Mahmoud WS. Medical Students’ Opinion Toward the Application of Complementary and Alternative Medicine in Healthcare. Saudi Journal of Medicine & Medical Sciences 2017;5:20–5. 10.4103/1658-631X.194255.

[46] Cutshall SM, Khalsa TK, Chon TY, Vitek SM, Clark SD, Blomberg DL, et al. Curricular Development and Implementation of a Longitudinal Integrative Medicine Education Experience for Trainees and Health-Care Professionals at an Academic Medical Center. Global Advances in Health and Medicine 2019;8:2164956119837489. 10.1177/2164956119837489.

[47] Verte LS. PROFILES - Principal Relevant Objectives and Framework for Integrative Learning and Education in Switzerland n.d. https://www.profilesmed.ch/epas/1-take-a-medical-history (accessed October 4, 2024).

[48] Lee MY, Benn R, Wimsatt L, Cornman J, Hedgecock J, Gerik S, et al. Integrating Complementary and Alternative Medicine Instruction into Health Professions Education: Organizational and Instructional Strategies. Academic Medicine 2007;82:939. 10.1097/ACM.0b013e318149ebf8.

